# Longitudinal symptom dynamics of COVID-19 infection in primary care

**DOI:** 10.1101/2020.07.13.20151795

**Authors:** Barak Mizrahi, Smadar Shilo, Hagai Rossman, Nir Kalkstein, Karni Marcus, Yael Barer, Ayya Keshet, Na’ama Shamir-Stein, Varda Shalev, Anat Ekka Zohar, Gabriel Chodick, Eran Segal

## Abstract

**Objective:** Data regarding the clinical characteristics of COVID-19 infection is rapidly accumulating. However, most studies thus far are based on hospitalized patients and lack longitudinal follow up. As the majority of COVID-19 cases are not hospitalized, prospective studies of symptoms in the population presenting to primary care are needed. Here, we assess the longitudinal dynamic of clinical symptoms in non-hospitalized individuals prior to and throughout the diagnosis of SARS-CoV-2 infection.

**Design:** Data on symptoms were extracted from electronic health records (EHR) consisting of both results of PCR tests and symptoms recorded by primary care physicians, and linked longitudinal self reported symptoms.

**Setting:** The second largest Health Maintenance Organization in Israel, Maccabi Health Services

**Participants:** From 1/3/2020 to 07/06/2020, information on symptoms from either surveys or primary care visits was available for 206,377 individuals, including 2,471 who tested positive for COVID-19.

**Main Outcomes:** Longitudinal prevalence of clinical symptoms in COVID-19 infection diagnosed by PCR testing for SARS-CoV-2 from nasopharyngeal swabs.

**Results:** In adults, the most prevalent symptoms recorded in EHR were cough (11.6%), fever (10.3%), and myalgia (7.7%) and the most prevalent self-reported symptoms were cough (21%), fatigue (19%) and rhinorrhea and/or nasal congestion (17%). In children, the most prevalent symptoms recorded in the EHR were fever (7%), cough (5.5%) and abdominal pain (2.4%). Emotional disturbances were documented in 15.9% of the positive adults and 4.2% of the children. Loss of taste and smell, either self-reported or documented by a physician, 3 weeks prior to testing, were the most discriminative symptoms in adults (OR =11.18 and OR=5.47 respectively). Additional symptoms included self reported headache (OR = 2.03) and fatigue (OR = 1.73) and a documentation of syncope, rhinorrhea (OR = 2.09 for both) and fever (OR= 1.62) by a physician. Mean time to recovery was 23.5 ± 9.9 days. Children had a significantly shorter disease duration (21.7 ± 8.8 days, p-value=0.01). Several symptoms, including fatigue, myalgia, runny nose and shortness of breath were reported weeks after recovery.

**Conclusions:** As the COVID-19 pandemic progresses rapidly worldwide, obtaining accurate information on symptoms and their progression is of essence. Our study shed light on the full clinical spectrum of symptoms experienced by infected individuals in primary care, and may alert physicians for the possibility of COVID-19 infection.

## Introduction

In December 2019 a cluster of severe respiratory disease from an unknown cause was identified in Wuhan, China. Soon after, the causative pathogen was identified as a novel coronavirus and was named the severe acute respiratory syndrome coronavirus 2 (SARS-CoV-2) ^1^. Since then, the virus has rapidly spread across China and worldwide, affecting thus far more than 12,000,000 confirmed patients and 500,000 deaths worldwide in above 200 Countries, areas or territories with confirmed cases, creating a major global health crisis ^2^.

As the virus spread rapidly across the globe, causing an increased number of infected individuals, reports on the clinical characteristics of the disease have started to emerge. Fever, cough, myalgia and fatigue were described as common symptoms of the disease. Less common symptoms included sputum production, headache, haemoptysis and diarrhea ^3–8^. Further on, anosmia and ageusia also emerged as prevalent and relatively discriminative symptoms of COVID-19 infection ^9–12^. As clinical data started to accumulate, reviews describing the potential cardiovascular ^13^, gastrointestinal ^14^, neurological ^15^ and cutaneous manifestations ^16^ of the disease were published.

The majority of studies published to date describing the clinical course of patients with COVID-19 infection were based on retrospective data of adult hospitalized patients^3–8^. This creates a knowledge gap, as infected people who only experience mild symptoms, do not necessarily seek clinical care. The clinical course of COVID-19 in children is less described, with more than 90% of all pediatric patients were previously described as asymptomatic, mild, or moderate ^17^, Information regarding the dynamic of symptoms throughout the disease course and their overall duration in both children and adults is still lacking.

In Israel, the first infection of COVID-19 was confirmed on February 21st 2020. In response, the Israeli Ministry of Health (MOH) employed a series of physical distancing measures in an attempt to mitigate the spread of the virus, which were later on gradually relieved, resulting in more than 25,000 diagnosed COVID-9 cases to date in Israel. Throughout the pandemic, testing policy in Israel has changed. Initially, only individuals who fulfilled both clinical criteria (symptoms of fever or respiratory symptoms) and epidemiological criteria (such as being in proximity to COVID-19 patients) were tested but further on, more indications for testing were added ^18^. Here, we analysed a unique dataset composed of electronic health records (EHR) from Maccabi Health Services (MHS), the second largest Health Maintenance Organization (HMO) in Israel which includes the results of SARS-CoV-2 PCR testing and primary care visits, and linked longitudinal self reported symptoms reported as part of a nationwide survey ^19^, to better understand the full clinical spectrum of symptoms experienced by adults and children infected with COVID-19.

## Methods

### Data

In this study, we utilized data originating from MHS. MHS is the second largest HMO currently active in Israel, representing a quarter of the Israeli population. It contains longitudinal data on over 2.3 million people since 1993, with annual attrition rate lower than 1%. The dataset included extensive demographic data, anthropometric measurements, clinic and hospital diagnoses, medication dispensed, and comprehensive laboratory data from a single central lab ^20^.

For adults, information on symptoms was obtained from two different data sources: symptoms documented in the EHR as ICD-9 codes and symptoms from a self reported survey. The survey was distributed by MHS from 01/04/2020 and throughout the COVID-19 pandemic to all adult members (age above 18 years of age) of MHS. The survey included questions relating to age, gender, prior medical conditions, smoking habits, self-reported symptoms and geographical location (see section 1 in the supplementary appendix). Each participant was asked to fill the survey once a day. The surveys were linked to the EHR by a unique identifier. For children, symptoms were extracted solely from the EHR, as the surveys were not distributed in this age group.

### Study Outcome

Patients with COVID-19 infection were identified as those having at least one record of a positive SARS-CoV-2 polymerase chain reaction (PCR) test in the MHS EHR. PCR tests for SARS-CoV-2 were obtained from nasopharyngeal swabs. Individuals negative to COVID-19 infection were considered as such if all their laboratory tests for SARS-CoV-2 were negative.

### Study Design and Population

To obtain information on symptoms in COVID-19 patients, we used two different data sources: symptoms recorded by physicians as ICD-9 codes and self reported symptoms from the survey (Fig 1).

**Figure 1.**
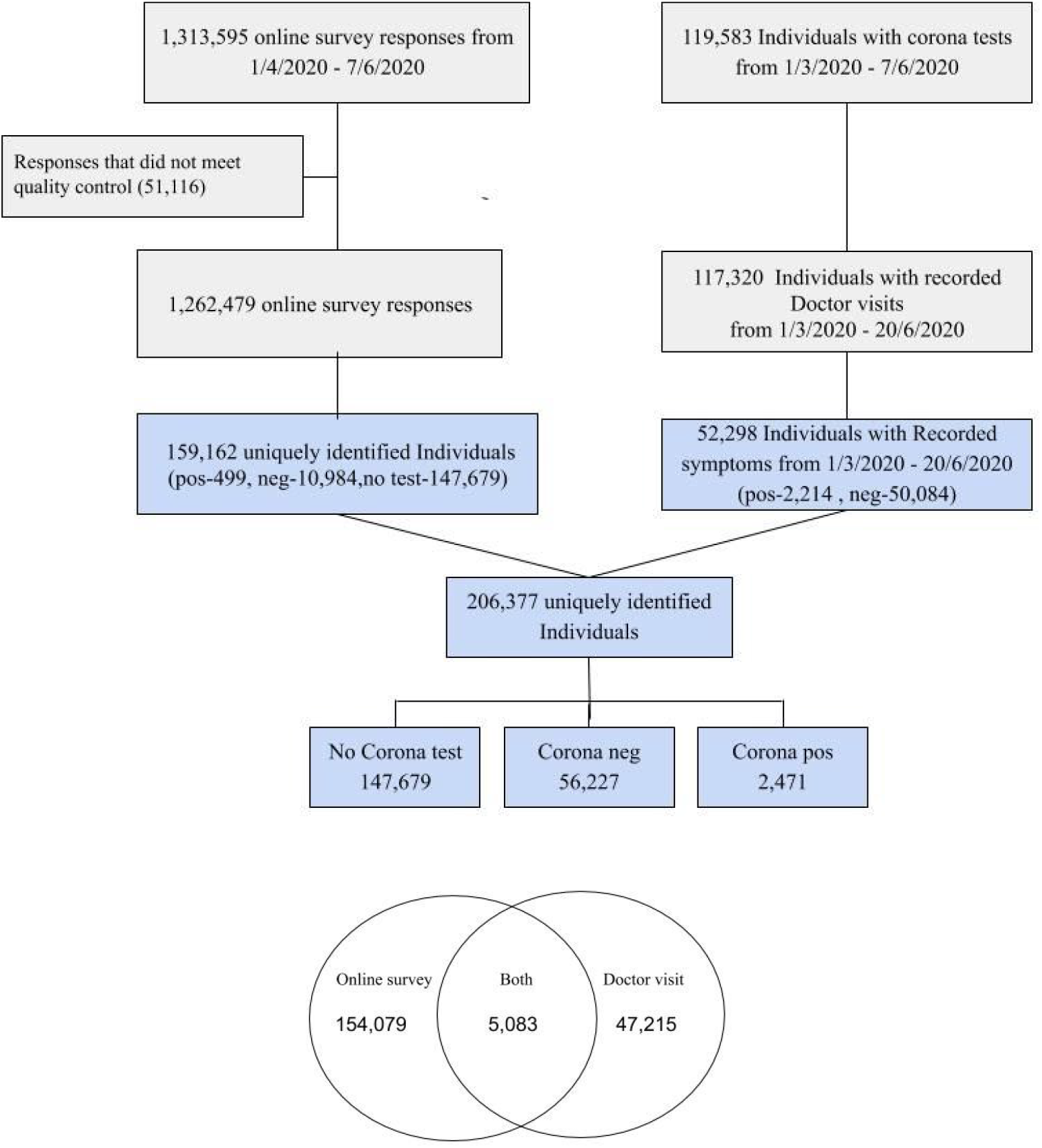
Cohort selection. **A:** A flow chart of cohort selection **B:** Venn diagram of the data sources from which information on symptoms were obtained. The number of individuals that had information on symptoms from the online survey, primary care visits recorded in the electronic health records and both data sources are presented.

First, we analyzed data of individuals in MHS, who had at least one PCR test for SARS-CoV-2 between 01/03/2020 and 07/06/2020, and had at least one primary care visit. During the COVID-19 pandemic, visits were held as in-person visits or telemedicine visits. Overall, 175,994 PCR tests for SARS-CoV-2, for 119,583 individuals, were performed during this time period. To capture diagnostic codes given after the date of the diagnostic PCR test, data from primary care visits were extracted from 01/03/2020 to 20/06/2020. During this time period, 117,230 individuals had a documented primary care visit. The characteristics of these individuals are described in Table 1. In order to distill symptoms that are prevalent in COVID19 cases, we first extracted all ICD-9 codes that were documented by a physician during the study period. From these, ICD-9 codes which represent symptoms that were previously described in COVID19 patients were analysed (see section 2 in the supplementary appendix).

**Table 1.**
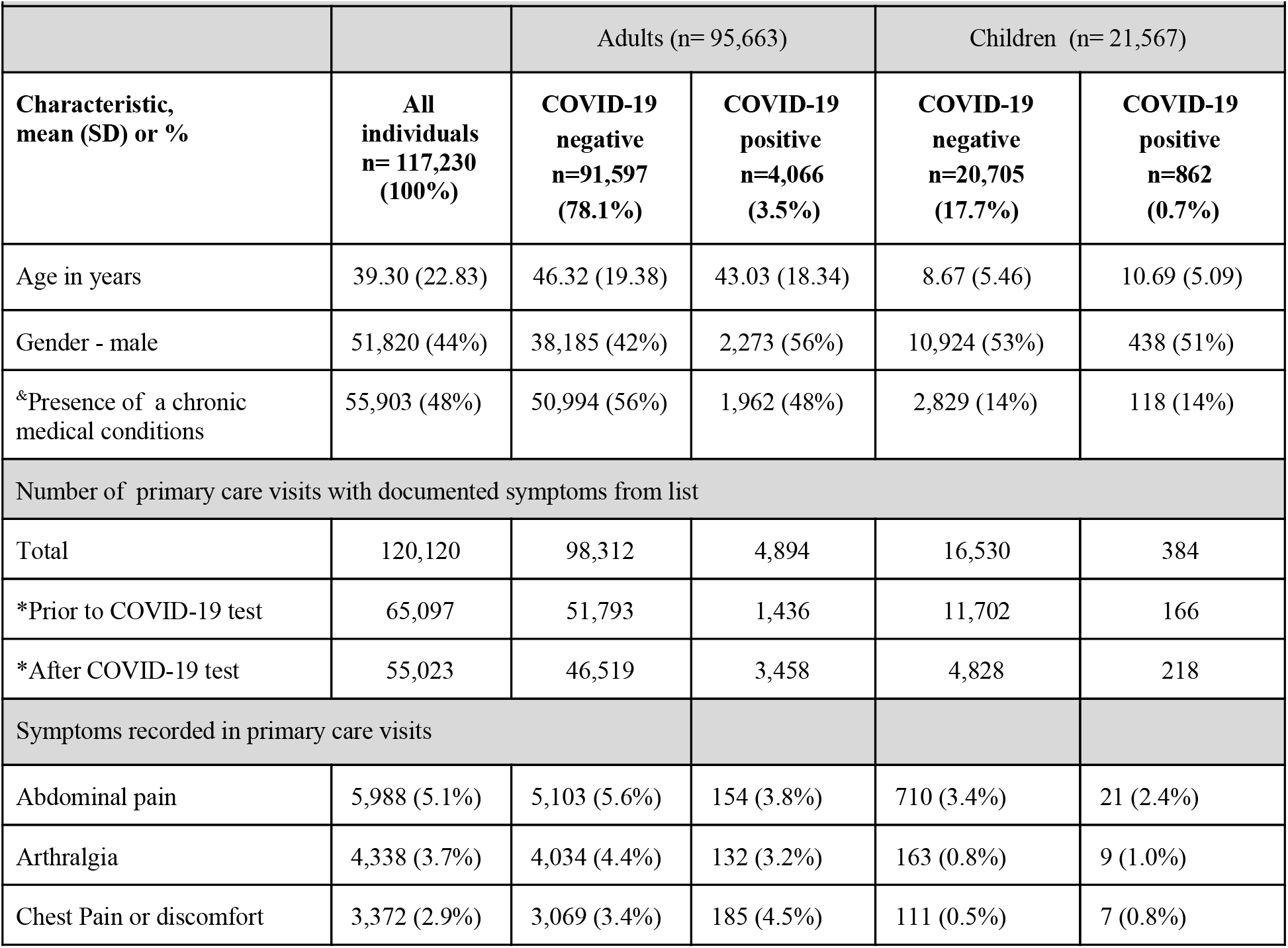
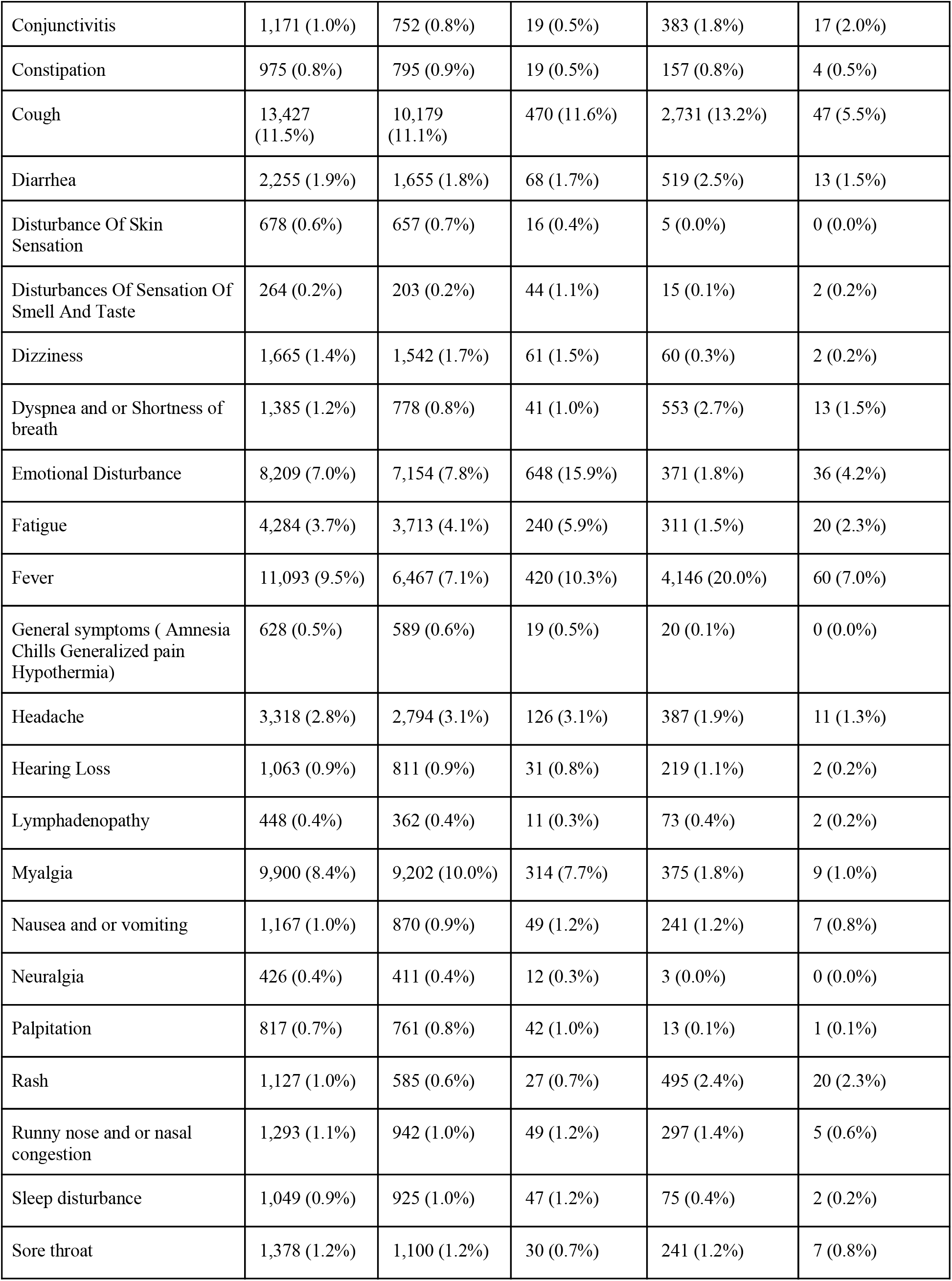
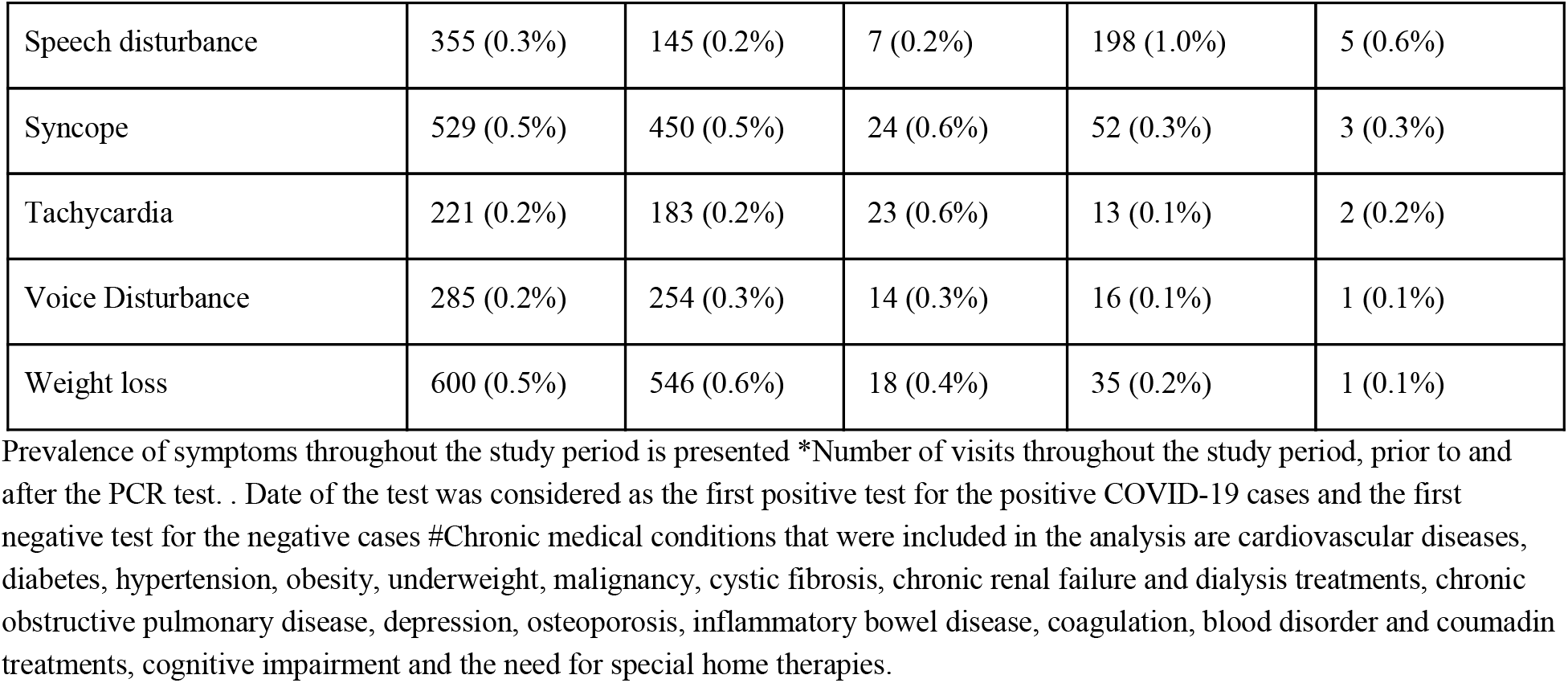
Baseline characteristics of the individuals with primary care visits.

|  |  |  |  |  |  |
| --- | --- | --- | --- | --- | --- |
|  |  | Adults (n= 95,663) |  | Children (n= 21,567) |  |
| **Characteristic, mean (SD) or %** | **All individuals n= 117,230 (100%)** | **COVID-19 negative n=91,597 (78.1%)** | **COVID-19 positive n=4,066 (3.5%)** | **COVID-19 negative n=20,705 (17.7%)** | **COVID-19 positive n=862 (0.7%)** |
| Age in years | 39.30 (22.83) | 46.32 (19.38) | 43.03 (18.34) | 8.67 (5.46) | 10.69 (5.09) |
| Gender - male | 51,820 (44%) | 38,185 (42%) | 2,273 (56%) | 10,924 (53%) | 438 (51%) |
| &Presence of a chronic medical conditions | 55,903 (48%) | 50,994 (56%) | 1,962 (48%) | 2,829 (14%) | 118 (14%) |
| Number of primary care visits with documented symptoms from list |  |  |  |  |  |
| Total | 120,120 | 98,312 | 4,894 | 16,530 | 384 |
| *Prior to COVID-19 test | 65,097 | 51,793 | 1,436 | 11,702 | 166 |
| *After COVID-19 test | 55,023 | 46,519 | 3,458 | 4,828 | 218 |
| Symptoms recorded in primary care visits |  |  |  |  |  |
| Abdominal pain | 5,988 (5.1%) | 5,103 (5.6%) | 154 (3.8%) | 710 (3.4%) | 21 (2.4%) |
| Arthralgia | 4,338 (3.7%) | 4,034 (4.4%) | 132 (3.2%) | 163 (0.8%) | 9 (1.0%) |
| Chest Pain or discomfort | 3,372 (2.9%) | 3,069 (3.4%) | 185 (4.5%) | 111 (0.5%) | 7 (0.8%) |
| Conjunctivitis | 1,171 (1.0%) | 752 (0.8%) | 19 (0.5%) | 383 (1.8%) | 17 (2.0%) |
| Constipation | 975 (0.8%) | 795 (0.9%) | 19 (0.5%) | 157 (0.8%) | 4 (0.5%) |
| Cough | 13,427 (11.5%) | 10,179 (11.1%) | 470 (11.6%) | 2,731 (13.2%) | 47 (5.5%) |
| Diarrhea | 2,255 (1.9%) | 1,655 (1.8%) | 68 (1.7%) | 519 (2.5%) | 13 (1.5%) |
| Disturbance Of Skin Sensation | 678 (0.6%) | 657 (0.7%) | 16 (0.4%) | 5 (0.0%) | 0 (0.0%) |
| Disturbances Of Sensation Of Smell And Taste | 264 (0.2%) | 203 (0.2%) | 44 (1.1%) | 15 (0.1%) | 2 (0.2%) |
| Dizziness | 1,665 (1.4%) | 1,542 (1.7%) | 61 (1.5%) | 60 (0.3%) | 2 (0.2%) |
| Dyspnea and or Shortness of breath | 1,385 (1.2%) | 778 (0.8%) | 41 (1.0%) | 553 (2.7%) | 13 (1.5%) |
| Emotional Disturbance | 8,209 (7.0%) | 7,154 (7.8%) | 648 (15.9%) | 371 (1.8%) | 36 (4.2%) |
| Fatigue | 4,284 (3.7%) | 3,713 (4.1%) | 240 (5.9%) | 311 (1.5%) | 20 (2.3%) |
| Fever | 11,093 (9.5%) | 6,467 (7.1%) | 420 (10.3%) | 4,146 (20.0%) | 60 (7.0%) |
| General symptoms ( Amnesia Chills Generalized pain Hypothermia) | 628 (0.5%) | 589 (0.6%) | 19 (0.5%) | 20 (0.1%) | 0 (0.0%) |
| Headache | 3,318 (2.8%) | 2,794 (3.1%) | 126 (3.1%) | 387 (1.9%) | 11 (1.3%) |
| Hearing Loss | 1,063 (0.9%) | 811 (0.9%) | 31 (0.8%) | 219 (1.1%) | 2 (0.2%) |
| Lymphadenopathy | 448 (0.4%) | 362 (0.4%) | 11 (0.3%) | 73 (0.4%) | 2 (0.2%) |
| Myalgia | 9,900 (8.4%) | 9,202 (10.0%) | 314 (7.7%) | 375 (1.8%) | 9 (1.0%) |
| Nausea and or vomiting | 1,167 (1.0%) | 870 (0.9%) | 49 (1.2%) | 241 (1.2%) | 7 (0.8%) |
| Neuralgia | 426 (0.4%) | 411 (0.4%) | 12 (0.3%) | 3 (0.0%) | 0 (0.0%) |
| Palpitation | 817 (0.7%) | 761 (0.8%) | 42 (1.0%) | 13 (0.1%) | 1 (0.1%) |
| Rash | 1,127 (1.0%) | 585 (0.6%) | 27 (0.7%) | 495 (2.4%) | 20 (2.3%) |
| Runny nose and or nasal congestion | 1,293 (1.1%) | 942 (1.0%) | 49 (1.2%) | 297 (1.4%) | 5 (0.6%) |
| Sleep disturbance | 1,049 (0.9%) | 925 (1.0%) | 47 (1.2%) | 75 (0.4%) | 2 (0.2%) |
| Sore throat | 1,378 (1.2%) | 1,100 (1.2%) | 30 (0.7%) | 241 (1.2%) | 7 (0.8%) |
| Speech disturbance | 355 (0.3%) | 145 (0.2%) | 7 (0.2%) | 198 (1.0%) | 5 (0.6%) |
| Syncope | 529 (0.5%) | 450 (0.5%) | 24 (0.6%) | 52 (0.3%) | 3 (0.3%) |
| Tachycardia | 221 (0.2%) | 183 (0.2%) | 23 (0.6%) | 13 (0.1%) | 2 (0.2%) |
| Voice Disturbance | 285 (0.2%) | 254 (0.3%) | 14 (0.3%) | 16 (0.1%) | 1 (0.1%) |
| Weight loss | 600 (0.5%) | 546 (0.6%) | 18 (0.4%) | 35 (0.2%) | 1 (0.1%) |

Prevalence of symptoms throughout the study period is presented \*Number of visits throughout the study period, prior to and after the PCR test. . Date of the test was considered as the first positive test for the positive COVID-19 cases and the first negative test for the negative cases #Chronic medical conditions that were included in the analysis are cardiovascular diseases, diabetes, hypertension, obesity, underweight, malignancy, cystic fibrosis, chronic renal failure and dialysis treatments, chronic obstructive pulmonary disease, depression, osteoporosis, inflammatory bowel disease, coagulation, blood disorder and coumadin treatments, cognitive impairment and the need for special home therapies.

Second, we analyzed data of individuals in MHS, who had at least one valid self-reported symptoms survey. Between 01/04/2020 to 7/06/2020, 1,313,595 surveys were filled for 181,798 individuals. From them 51,116 (3.9%) surveys were not fully completed by participants and were excluded from the analysis. A total of 1,262,479 surveys for 159,162 adults were included in the analyses. The characteristics of these individuals are described in Table 2. Self reported fever was defined as a reported body temperature above 38°C. Chronic medical disease were defined as one of the following: cardiovascular diseases, diabetes, hypertension, obesity, underweight, malignancy, cystic fibrosis, chronic renal failure and dialysis treatments, chronic obstructive pulmonary disease, depression, osteoporosis, inflammatory bowel disease, coagulation, blood disorder and coumadin treatments, cognitive impairment and the need for special home therapies ^21^. 5,083 individuals in MHS who performed a PCR test for SARS-CoV-2, had both a documented primary care visit with recorded symptoms and filed a self-reported symptoms survey. These individuals were included in both analyses. The medical charts and self reported symptoms of individual cases of COVID-19 patients were reviewed by two physicians.

**Table 2.** Baseline characteristics of the survey responders.

| **Table 2. Baseline characteristics of the survey responders** |  |  |  |  |
| --- | --- | --- | --- | --- |
| **Characteristic, mean (SD) or %** | **All individuals**<br>**n= 159,162**<br>**(100%)** | **Individuals**<br>**not tested for**<br>**COVID-19**<br>**N = 147,679**<br>**(92.78%)** | **COVID-19**<br>**negative**<br>**n= 10,984**<br>**(6.9%)** | **COVID-19**<br>**positive**<br>**n= 499**<br>**(0.31%)** |
| Age in years | 46.62 (16.94) | 46.84 (16.95) | 43.82 (16.45) | 42.15 (16.61) |
| Gender - male | 66,777 (42%) | 62,461 (42%) | 4,085 (37%) | 231 (46%) |
| &Presence of a chronic medical conditions |  |  | 5,789 (53%) | 231 (46%) |
| Healthcare workers | 9,934 (6%) | 8,279 (6%) | 1,602 (15%) | 53 (11%) |
| **Self reported surveys** |  |  |  |  |
| Total | 1,262,479 | 1,187,523 | 72,928 | 2,028 |
| *Prior to COVID-19 test |  |  | 36,169 | 371 |
| *After COVID-19 test |  |  | 36,759 | 1,657 |
| Per individual<br>Mean (SD)<br>Median(IQR) | 7.93 (9.96)<br>3 (1-11) | 8.04 (10.05)<br>3 (1-11) | 6.64 (8.72)<br>2 (1-8) | 4.06 (5.98)<br>2 (1-4) |
| **Symptoms** |  |  |  |  |
| Fever (body temperature above 38 °C) | 863 (1%) | 602 (0%) | 250 (2%) | 11(2%) |
| Body temperature between 37.5-37.9 °C | 1,868 (1%) | 1,566 (1%) | 294 (3%) | 8(2%) |
| Body temperature below 37.4 °C | 100,719 (63%) | 93,021 (63%) | 7,350 (67%) | 348 (70%) |
| Fatigue | 12,630 (8%) | 10,745 (7%) | 1,792 (16%) | 93(19%) |
| Nausea and/or vomiting | 3,793 (2%) | 3,290 (2%) | 488 (4%) | 15(3%) |
| Confusion | 1,364 (1%) | 1,143 (1%) | 206 (2%) | 15(3%) |
| Rhinorrhea and/or nasal congestion | 25,828 (16%) | 23,205 (16%) | 2,536 (23%) | 87(17%) |
| Sore throat | 13,684 (9%) | 11,766 (8%) | 1,870 (17%) | 48(10%) |
| headache | 19,818 (12%) | 17,585 (12%) | 2,152 (20%) | 81(16%) |
| Myalgia | 9,011 (6%) | 7,766 (5%) | 1,192 (11%) | 53(11%) |
| Shortness of breath | 4,248 (3%) | 3,515 (2%) | 709 (6%) | 24(5%) |
| Feel well | 128,633 (81%) | 120,424 (82%) | 7,860 (72%) | 349(70%) |
| Other symptoms | 11,066 (7%) | 10,015 (7%) | 1,004 (9%) | 47(9%) |
| Diarrhea | 6,889 (4%) | 6,060 (4%) | 804 (7%) | 25(5%) |
| Cough | 20,162 (13%) | 17,374 (12%) | 2,682 (24%) | 106(21%) |
| Wet cough | 11,509 (7%) | 9,978 (7%) | 1,482 (13%) | 49(10%) |
| Dry cough | 11,340 (7%) | 9,586 (6%) | 1,676 (15%) | 78(16%) |
| Loss of taste or smell | 1,209 (1%) | 951 (1%) | 209 (2%) | 49(10%) |
| Chills | 2,430 (2%) | 1,951 (1%) | 467 (4%) | 12(2%) |
| **Isolation status** |  |  |  |  |
| International travel | 841 (0.53%) | 638 (0.43%) | 198 (1.80%) | 5 (1.00%) |
| Contact with a positive COVID-19 case | 1429 (0.90%) | 815 (0.55%) | 570 (5.19%) | 44 (8.82%) |
| Experiencing disease symptoms | 1240 (0.78%) | 597 (0.40%) | 621 (5.65%) | 22 (4.41%) |
| personal preference | 11040 (6.94%) | 10281 (6.96%) | 742 (6.76%) | 17 (3.41%) |
| Other reason | 3777 (2.37%) | 3312 (2.24%) | 458 (4.17%) | 7 (1.40%) |

Prevalence of symptoms throughout the study period is presented \*Number of filled surveys throughout the study period, prior to and after the PCR test. Date of the test was considered as the first positive test for the positive COVID-19 cases and the first negative test for the negative cases. #Chronic medical conditions that were included in the analysis are cardiovascular diseases, diabetes, hypertension, obesity, underweight, malignancy, cystic fibrosis, chronic renal failure and dialysis treatments, chronic obstructive pulmonary disease, depression, osteoporosis, inflammatory bowel disease, coagulation, blood disorder and coumadin treatments, cognitive impairment and the need for special home therapies.

### Statistical analysis

To analyse the dynamic of symptoms in COVID-19 cases over time, we plotted the percentages of reported symptoms across different days relative to the time of COVID-19 test. This was considered to be the first positive PCR test result for COVID-19 cases and the first negative PCR test result of negative COVID-19 cases. In addition, for positive COVID-19 cases, we analysed the prevalence of symptoms relative to the recovery date, which was considered as the date in which a second consecutive negative PCR test result for COVID-19 was recorded. These analyses were done separately for symptoms recorded in primary care visits and those which were self reported by participants through a survey. To evaluate the significance of differences in disease duration, a Mann whitney test was used.

## Results

From 1/3/2020 to 07/06/2020, information on symptoms was available for 206,377 individuals (see Methods, Fig 1). Information on symptoms was obtained from two sources: 117,230 individuals performed a PCR test for SARS-CoV-2 and had a record of primary care visit (Table 1). Of them, 52,298 had a primary care visit with documented symptoms (2,214 positive, 50,084 negative). 159,162 individuals (499 positive, 10,984 negative and 147,679 not tested) filled at least one-self reported symptoms survey (Table 2). 5,083 individuals had information on symptoms from both sources. A snapshot of the detailed clinical course for one individual with COVID-19 infection in our cohort is presented as an example in Figure 3.

Altogether, information on symptoms was available for 2,471 individuals defined as positive COVID-19 cases, 56,227 defined as negative and 147,679 individuals who had no record of a PCR test for SARS-CoV-2. Among the positive cases, and throughout the study period, 508 (20.6%) individuals were hospitalized, including 51 (2.1%) individuals that were admitted to the Intensive care unit (ICU) and 16 (0.6 %) patients died.

### Clinical manifestations of COVID-19 infection in adults

Overall, the most prevalent symptoms recorded in primary care visits throughout the study period in adult COVID-19 cases were cough (11.6%), Fever (10.3%), Myalgia (7.7%) and Fatigue (5.9%). Emotional disturbance, including anxiety and depression, were common (15.9%). The most prevalent self-reported symptoms were cough (21%), fatigue (19%), rhinorrhea and/or nasal congestion (17%), headache (16%) and myalgia (11%) (tables 1 & 2). Of note, in order to obtain the full clinical picture, responders to the survey were asked whether they experience additional symptoms and were given an option to report these in length and in a free text format. Only few of the positive COVID-19 cases reported additional symptoms and those were mainly reported before disease onset. These included: abdominal pain, chest discomfort/chest pain, loss of appetite, bitter taste and chills.

### Clinical manifestations of COVID-19 infection in children

A total of 21,567 children were included in the analysis. Of them, 862 (4%) were positive COVID-19 cases (mean age of 10.69± 5.09 years old) and 20,705 (96%) negative (mean age of 8.67±5.46 years old). The percentage of positive cases from all individuals tested was similar between children and adults (4% versus 4.25% respectively). Data on clinical symptoms of these children was obtained solely from the MHS EHR as the survey was not distributed at this age group. Conjunctivitis, rash, sore throat, dyspnea and/or shortness of breath and speech disturbance, had a higher prevalence in children who were positive to COVID-19 compared to positive adults. Overall, the most prevalent symptoms recorded in primary care visits throughout the study period were different than the adults’ cases, and included fever (7%), cough (5.5%), abdominal pain (2.4%) and fatigue (2.3%). Emotional disturbance, including anxiety and depression, were less documented in children compared to adults, but were present in 4.2% of positive cases. Most symptoms were rare, and were present in less than 1% of the patients.

### Dynamics of symptoms in COVID-19 patients

We next analysed the dynamics of symptoms reported by individuals with confirmed COVID-19 infection compared to individuals with negative COVID-19 tests in time. To that end, we made use of the rich individual level data obtained from both of the data sources we had in our possession: the self-reported symptoms and the symptoms which were recorded in the EHR during primary care visits. These data were analysed separately (see Methods).

A longitudinal analysis of symptoms on the entire cohort from both data sources and in regard to the time of test and time of recovery revealed different patterns in time for different symptoms (Fig 2). Interestingly, there is an increase in loss of taste and smell sensation in COVID-19 patients, as compared to negative cases, which is apparent both in the self-reported surveys, and in a smaller magnitude also in primary care visits. The prevalence of these symptoms start increasing as early as 3 weeks prior to time of diagnosis, decreasing up to approximately a week prior to recovery date.

**Figure 2.**
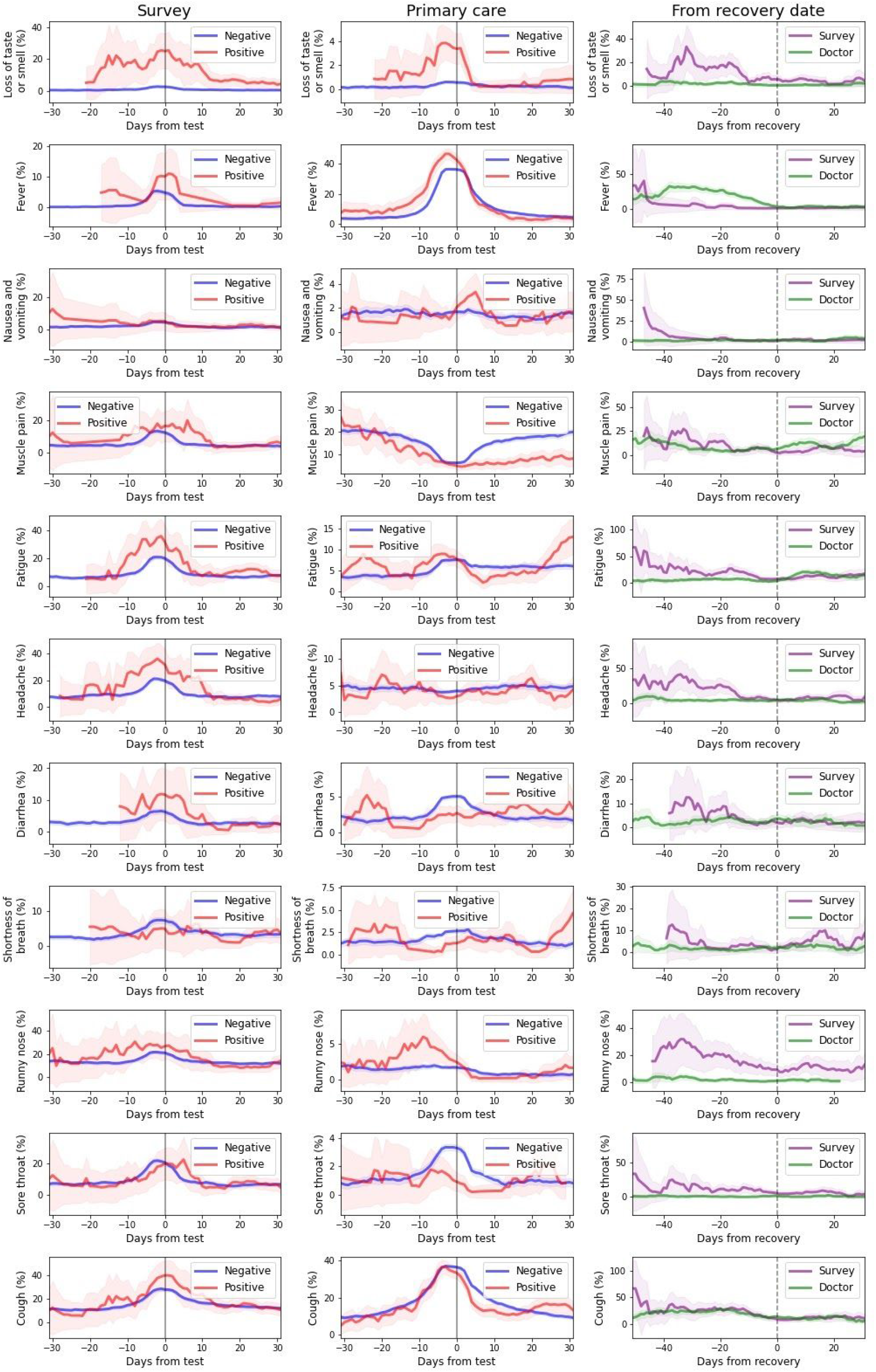
Dynamics of symptoms in COVID-19 patients. each row represent the prevalence of a symptom in our cohort analysed by time relative to diagnosis day from **A:** Survey of self reported symptoms and **B:** Primary care visits in positive COVID19 cases (red) versus negative (blue) **C** Prevalence of symptoms in our cohort relative to time of recovery by surveys of self reported symptoms (purple) and primary care visits (green). Each time point is calculated by taking a 1 week window (±3 days from day). Note y-axis scale is different for each panel.

**Figure 3.**
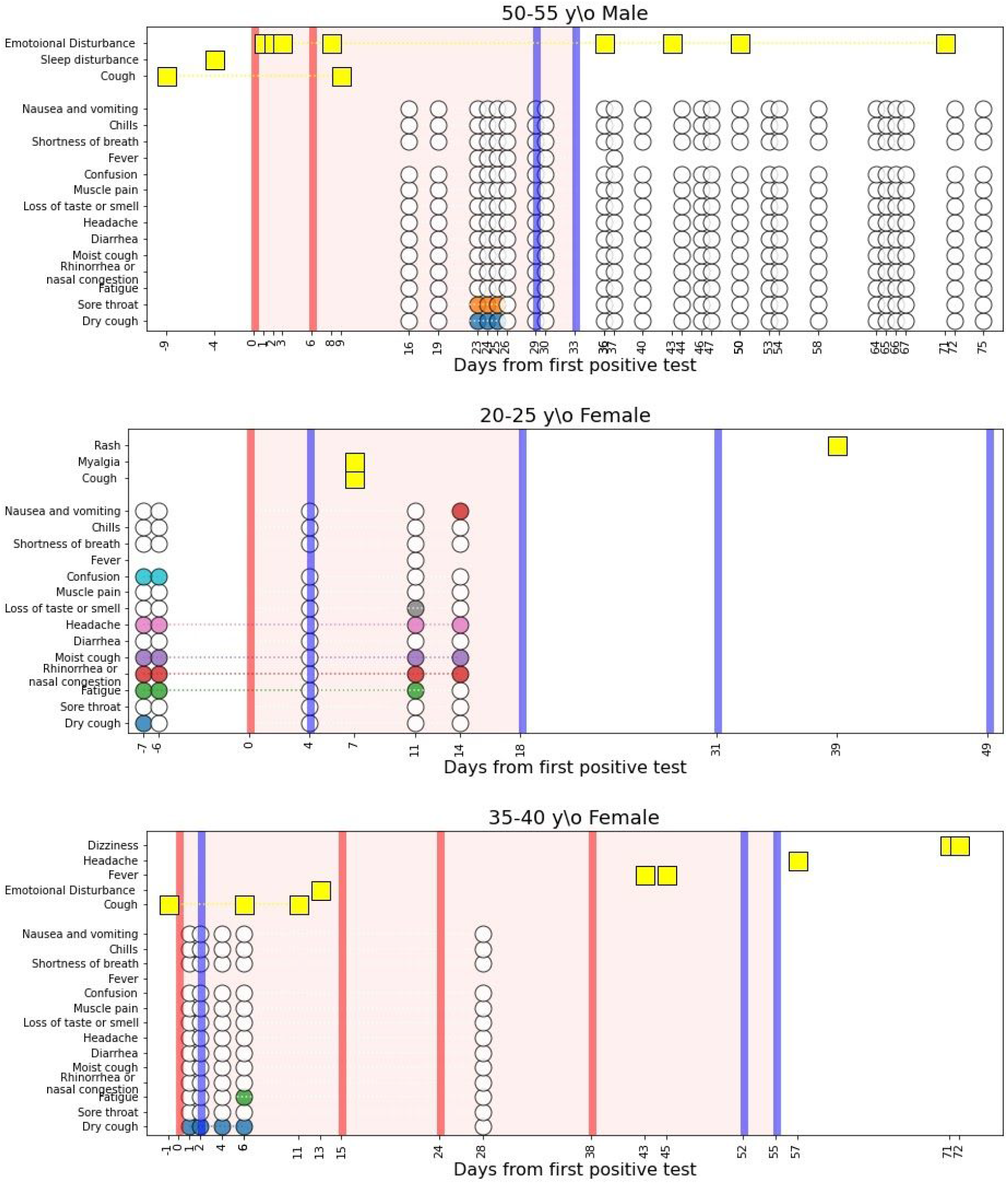
Individual cases. Longitudinal dynamics of symptoms in infected individuals. 3 examples are given. Yellow rectangles represent symptoms recorded by a physician at a primary care visit. Colored circles represent symptoms self-reported by the individual throughout the survey. White circles represent self-report of not experiencing a symptom through the survey. Red and blue vertical lines indicate a positive or negative PCR test for SARS-CoV-2 respectively. The area marked in light red indicates the period of time an individual was considered as infected with COVID-19.

**Figure 4.**
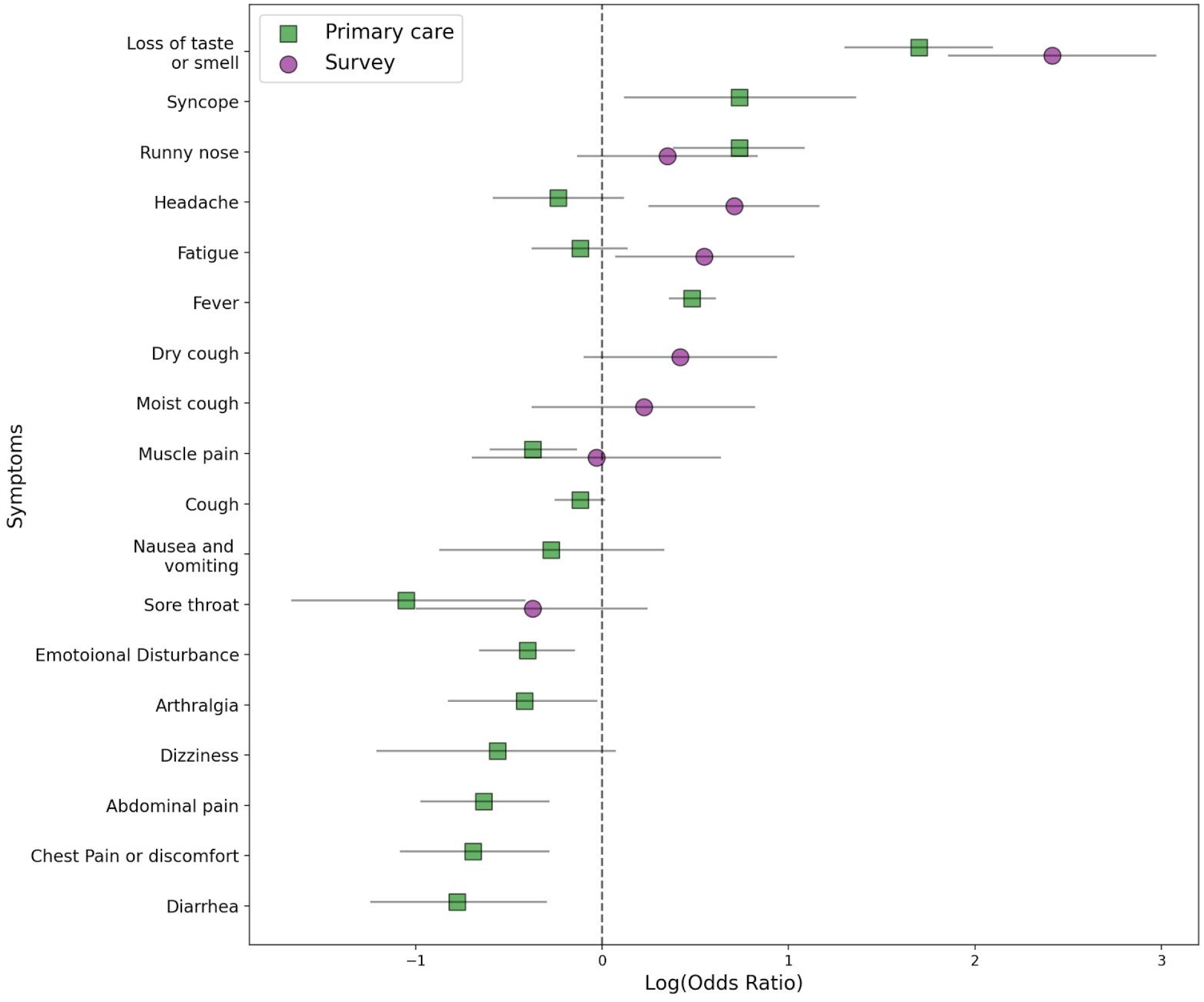
Odds ratio analysis of symptoms prior to COVID-19 test. Log odds ratio calculated by the prevalence of symptoms 21 days prior to COVID-19 test for adults.. Green squares are Log odds ratios calculated from primary care visits. Purple circles are calculated from symptom surveys. Grey lines indicate 95% confidence intervals.

As fever was one of the main criteria to fulfill in order to be applicable for COVID19 test in Israel in the majority of time during the COVID-19 pandemic, it is not surprising that we observe an increase in this symptom in both positive and negative cases with a peak just before the time of testing, and that this symptom is more evident in primary care visits. Nonetheless, the percentage of fever in positive cases is still higher than the negative cases, both in the survey and in primary care visits. Similarly, cough was also a part of the criteria for COVID19 testing and also had a peak in prevalence just before the time of testing for both positive and negative cases.

Fatigue was also self-reported by participants in a higher prevalence around the time of testing in both positive and negative cases, but with a higher prevalence in the positive cases. The dynamics of Sore throat and diarrhea was similar in positive and negative cases with the maximal prevalence reported around the diagnostic test. Higher prevalence of these symptoms was documented by physicians in negative cases at time of diagnosis. Self reported headache was more prevalent in positive cases, mainifistated in up to 30% of patients prior to the day of diagnosis, and gradually decreasing up to 2 weeks before recovery when it is diminished. The dynamics of additional symptoms which were reported by individuals in our cohort or recorded by their physicians is presented in Figure 2.

### Clinical manifestations of COVID-19 infection after recovery

Data on symptoms after recovery were obtained from both data sources. From 2,214 positive cases who had a record of primary care visit with symptoms, 1,818 recovered throughout the study period, and from them, 909 had a primary care visit after recovery, with average follow up time of 31.4 ± 20.1 days after recovery. From 499 positive cases who filled at least one-self reported symptoms survey, 442 recovered, 278 of them have filled a survey after recovery with an average follow up time of 17.6 ± 14.8 days. Long duration of symptoms, specifically fatigue, myalgia, runny nose and shortness of breath was observed weeks after recovery.

### Variability in the clinical course of COVID-19 infection

After analysing the general dynamics of symptoms, we further looked at the variability of the clinical course in different individuals in the cohort. A high variability in the clinical course was observed by two physicians who reviewed the medical charts and surveys of COVID-19 cases. Clinical spectrum was ranging from mild and short disease to a prolonged course, lasting weeks after the recovery. Several examples are shown in Figure 3. Next, we analysed the duration of disease, calculated by the number of days from the date of the first positive PCR test to recovery date. This analysis revealed a mean duration of 23.5 ± 9.9 days (n=2,045), thus showing a high variability of disease duration. Younger age was significantly associated with a shorter disease: children had an average duration of 21.7 ± 8.8 (n=177) to recovery compared to adults, who had 23.7 ± 9.9 (n=1,868) days to recovery (p-value = 0.01). Gender did not affect time to recovery : females had an average duration of 23.8 ± 9.9 days (n=952) to recovery compared to males 23.3 ± 9.8 (n=1093) (p-value = 0.157). Next, we analysed whether the presence of specific symptoms was associated with different time to recovery. This analysis revealed that for example, individuals with loss of smell and taste tend to have a shorter time to recovery compared to those experiencing shortness of breath, possibly due to the fact that the latter represents a disease in higher severity (see section 3 in the supplementary appendix, Fig S1).

### Odds-ratio analysis

In order to distill the symptoms that can assist physicians in identifying COVID-19 patients prior to diagnosis, we next analysed the association between symptoms experienced by individuals in the 3 weeks prior to the time of testing and COVID-19 diagnosis and were present in at least 10 positive cases (Fig 4). This analysis was done separately for children and adults. For adults, loss of taste and smell, either self-reported or documented by physician, were the symptoms which were most associated with a positive diagnosis (odds ratio (OR) = 11.18; 95% confidence interval (CI) 6.43-19.44 and OR=5.47 (3.69-8.09) for self-reported and primary care visit documentation respectively).

Other self reported symptoms that were significantly associated with COVID-19 diagnosis were fatigue (OR = 1.73(1.08-2.79)) and headache (OR = 2.03 (1.29-3.19)). Syncope (OR = 2.09 (1.13-3.88)), runny nose (OR= 2.09 (1.47-2.95)) and fever (OR= 1.62 (1.44-1.83)) documented by a physician in a primary care visit were also significantly associated with a positive diagnosis. Self reported confusion was also associated with positive cases but was reported by only 5 individuals in the 3 weeks prior to the diagnostic test (OR =4.02 (1.58-10.21)).

In contrast, a physician documentation of diarrhea, sore throat, abdominal or chest pain and Arthralgia were significantly associated with a negative COVID-19 cases (OR of 0.46 (0.29-0.74), 0.35 (0.19-0.66), 0.53 (0.38-0.75), 0.5 (0.34-0.75) and 0.66 (0.44-0.97) respectively). Results on self reports for these symptoms were only partially available and in some cases revealed an opposite trend, although not statistically significant.

In children, sample size was smaller and information on symptoms was only available from the EHR. Only 3 symptoms were experienced in the 3 weeks prior to the diagnostic test by more than 10 positive individuals: cough, fever and fatigue. Fever (OR = 0.3 (0.22-0.42) and cough (OR=0.4 (0.28-0.59) were associated with negative COVID-19 cases. Analysis of the less prevalent symptoms revealed that syncope and loss of taste and smell had the highest OR for COVID-19 infection (2.45 for both) but were not statistically significant while several symptoms were negatively associated with COVID-19, including diarrhea (OR = 0.17(0.04-0.7) and abdominal pain (OR= 0.1(0.01-0.69). Full results of OR analyses are presented in section 4 of the Supplementary appendix.

## Discussion

In this study, we leveraged a unique dataset that includes EHR from the second largest HMO in Israel, and linked longitudinal self reported symptoms surveys collected prior to and after COVID-19 testing. These two sources of information enable us to comprehensively capture data on symptoms of mostly mild COVID-19 cases from two different perspectives: the reports of the patient themself, and the reports of their physician. As previously reported, we observed a high variability in the clinical course of COVID-19 patients. Recovery time was highly variable, with a mean duration of 23.5 ± 9.9 days, and was significantly shorter in children (p-value = 0.01).

Temporal dynamics of self-reported and documented symptoms revealed different patterns and long duration of symptoms, specifically fatigue, myalgia, runny nose and shortness of breath was observed weeks after recovery. Previous studies which were mostly based on hospitalized patients have marked fever (90% or more), cough (around 75%), and dyspnea (up to 50%) as well as gastrointestinal symptoms in a small subset of patients to be the main clinical manifestations of COVID-19 ^22^. In our cohort, consisting of primarily non-hospitalized patients with mild disease, the most prevalent symptoms in adults recorded in EHR were cough (11.6%), fever (10.3%), myalgia (7.7%) and fatigue (5.9%) and the most prevalent self-reported symptoms were cough (21%), fatigue (19%) and rhinorrhea and/or nasal congestion (17%). In children, the most prevalent symptoms recorded in the EHR were fever (7%), cough (5.5%), abdominal pain (2.4%) and fatigue (2.3%).

Disturbances of the sensation of smell and taste was documented by physicians in 1.1% of the adult patient and 0.2% of the children but was self reported in 10% of the cases. Although these symptoms were not common, they emerged in our study as having the highest OR for COVID-19 diagnosis when appearing 3 weeks prior to COVID-19 tests in both adults and children (OR= 11.18 and OR=5.47 as self-reported or documented by physician for adults, and OR= 2.45 for children). In children, the result was not statistically significant, possibly due to a small sample size. Although anosmia and ageusia were initially less described as COVID-19 symptoms ^3,6^, and were not part of the Israeli testing policy throughout the study period, they further emerged as the most predictive symptoms for COVID-19, in this study and others ^9,11^, and health organisations worldwide have gradually added it to the list of COVID-19 related symptoms including the USA CDC ^23^, the UK ^24^ and Israel ^25^.

Other significant self reported symptoms which were reported by at least 10 positive COVID-19 cases in the 3 weeks prior to testing and are still not included in formal testing policies were headache (OR = 2.03) and fatigue (OR = 1.73). These symptoms are not specific to COVID-19, but may potentially help discriminate between positive and negative cases. Interestingly, syncope documented by a physician 3 weeks prior to diagnosis was significantly higher in positive cases in adults (OR = 2.09) and had a high OR but not statistically significant in children (OR = 2.45). Nonetheless, the overall prevalence of this symptom was very low in the cohort (0.6% and 0.3% of positive adults and children respectively) and reports on syncope as a COVID-19 related symptoms thus far were sparse ^26^. Therefore more studies are needed in order to determine if this symptom is part of COVID-19 clinical sequelae.

In children, several symptoms, including some which are part of the Isreali testing guidelines, such as cough (OR=0.4) and fever (OR = 0.3) were associated with negative cases, possibly due to the fact that many other infectious disease which are characterized by these symptoms were more prevalent in this age group during this time period.

Emotional disturbance, including anxiety and depression, were documented in 15.9% of the positive adults and 4.2% of the children, and appeared both prior to and after diagnosis. This emphasizes the fact that as the pandemic increases the risk for psychiatric illness, and in addition to medical care, health care providers have to closely monitor the psychosocial needs of their patients ^27^. An example is the establishment of new guidelines for emergency psychological crisis intervention initiated for people affected with COVID-19 by the National Health Commission of China ^28^.

Our study has several strengths. First, the integration of information on symptoms from two different data sources allowed us to comprehensively explore the clinical course of mild COVID-19 cases. As most of the studies on the clinical characteristics of COVID-19 infection thus far rely mostly on hospitalized patients, this information unreveal the heterogeneous clinical spectrum of symptoms experienced by COVID-19 infected individuals in the community setting, and may help physicians to identify individuals at risk. Second, data on the presence of clinical symptoms was available for 982 positive cases from either EHR or surveys up to 21 days prior to COVID-19 diagnosis, thus allowing us to capture symptoms prior to performing the diagnostic test in a relatively high percentage of the cohort (40%). This allowed us to prospectively analyse the dynamic of symptoms throughout the disease course, and not only after the diagnosis was already made. Third, COVID-19 cases were identified directly by a documented record of a positive PCR test in the EHR and were not based on self-reported diagnosis. Moreover, in order to check the validity of our survey, we directly compared self-reported answers on COVID-19 diagnoses in our survey to the documented test in the medical chart and found a very high agreement between the two sources. For example, from those who reported not diagnosed with COVID-19 in the survey, only 0.03% had a record of a positive COVID-19 test in the EHR thus further validating our survey.

Our study also has several limitations. First, the testing policy for COVID-19 in Israel has changed throughout the study period ^18^. In the majority of the time, individuals had to present with fever or respiratory symptoms, as well as an appropriate epidemiological context, in order to be tested. This may have a strong effect on the prevalence of these symptoms in individuals who were tested compared to those who were not. In addition, both of our data sources may introduce biases to the data and therefore were analysed separately by us. Data which is based on the voluntary self reported symptoms of participants is bound to suffer from selection bias. The fact that the survey was distributed to all the members of MHS by the HMO itself may decrease this bias. Data originating from EHR may also suffer from biases related to processes within the healthcare system ^29^ and a bias toward patients with more severe conditions.

In conclusion, this study we analysed the clinical course of outpatients COVID-19 cases. The study provides additional information on the natural history of mostly mild cases of COVID-19 and may alert physicians for the possibility of the infection and direct the need for testing and self isolation

## Data availability statement

The data that support the findings of this study originate from Maccabi Health Services. Restrictions apply to the availability of these data and they are therefore not publicly available. Due to restrictions, these data can be accessed only by request to the authors and/or Maccabi Health Services.

## Code availability statement

Analysis code is available at https://github.com/barakm-ki/Simptoms-dynamics-of-COVID-19-infection though it is tailored to the data and the fields of the Maccabi Health Services database.

## Ethics Declarations

The study protocol was approved by Maccabi Health Services’ institutional review board (0024-20-MHS). Informed consent was waived by the IRB, as all identifying details of the participants were removed before the computational analysis.

## Competing Interests Statement

The authors declare no competing interests.

## Authors contribution

B.M., S.S. and H.R conceived the project, designed and conducted the analyses, interpreted the results and wrote the manuscript. N.K conceived and directed the project, designed and conducted the analysis, K.M. designed and conducted the analyses, interpreted the results and wrote the manuscript. N.S.S & A.E.Z. provided and interpreted data. A.K. interpreted the data. Y.B, V.S. G.C. directed the project as well as provided the data, E.S. conceived the project, designed and conducted the analyses, interpreted the results and supervised the project and analyses.

## Acknowledgments

We thank the following for their contributions to our efforts: Tomer Meir, Amir Gavrieli, Tal Karady, Anastasia Godneva, Saar Shoer, Amit Lavon,, Dimitry Kolobkov, Iris Kalka, Ori Cohen, Pini Akiva, Chen Yanover, Guy Amit, Irena Girshovitz, Esma Herzel, Brosh Yinon.

## Supplementary appendix

1. COVID-19 survey
2. Symptoms extraction from EHR
3. Symptoms and time of recovery analysis
4. Odds ratio analysis

#### Section 1: COVID-19 survey

Age:_____

Gender:

□ Male
□ Female

I am:

□ Feeling well
□ Not feeling well

Are you experiencing any of the following symptoms?

Cough

□ Dry cough (no sputum)
□ Wet cough (with sputum)
□ Fatigue
□ Muscle pain
□ Shortness of breath
□ Rhinorrhea (runny nose) and/or nasal congestion
□ Diarrea
□ Nausea and/or vomiting
□ Sore throat
□ Headache
□ Chills
□ Confusion
□ Loss of taste or smell
□ Other symptoms-_____

I am currently:

□ Not in isolation
□ In isolation (including from family members, staying in a separate room) from the date of ______ due to:
  □ A recent international travel
  □ A contact with an individual who was infected with coronavirus
  □ Experiencing disease symptoms
  □ Voluntary isolation
□ I have a confirmed infection with COVID-19 (by a lab test) and currently:
  □ In home isolation
  □ Staying in a hotel
  □ Hospitalized in a hospital
  □ I recovered from COVID-19 infection

Cigarette smoking habits:

□ I currently smoke
□ I used to smoke and stopped more than 5 years ago
□ I used to smoke and stopped less than 5 years ago
□ I have never smoked

What is your current body temperature?

□ I did not measure my temperature in the last 24 hours
□ I measured my temperature (degrees celsius) and the highest value was :
  □ Below 36 °C
  □ 36-36.9 °C
  □ 37-37.4 °C
  □ 37.5-37.9 °C
  □ 38-38.4 °C
  □ 38.5-38.9 °C
  □ 39-39.9 °C
  □ Above 40 °C

How many individuals have you been in contact with in the last 24 hours? (within approximately 2 meters (6 ft 7 in) for more than 15 minutes)

Adults (age above18 years old______)

Children (age below 18 years old_______)

Do you work outside home?

□ Yes-Did you meet with more than 10 people a day at work in the last two weeks ? Yes/No
□ No

Do you, or anyone of your household members is a part of a medical team, actively treating patients?

□ Yes
□ No

#### Section 2: Extracting symptoms from EHR

Symptoms were extracted from the EHR using the relevant ICD-9 codes

| Symptoms group | ICD-9 Codes |
| --- | --- |
| Disturbances Of Sensation Of Smell And Taste | 781.1 - Disturbances Of Sensation Of Smell And Taste Anosmia; Parageusia; Parosmia |
| Emotional Disturbance | 309.1 - Adjustment Reaction With Prolonged Depressive Reaction |
|  | 309.8 - Other Specified Adjustment Reactions |
|  | 308.0 - Predominant Disturbance Of Emotions {Anxiety} {Emotional crisis} {Panic state} as acute reaction to exceptional [gross] stress |
|  | 799.90 - Diagnosis Or Condition Deferred On Axis 1 |
|  | 300.00 - Anxiety State, Unspecified Anxiety: neurosis, reaction, state (neurotic); Atypical anxiety disorder |
|  | 309.9 - Unspecified Adjustment Reaction Adaptation reaction NOS; Adjustment reaction NOS |
|  | 309.0 - Adjustment Reaction With Brief Depressive Reaction Adjustment disorder with depressed mood; Grief reaction |
|  | 300.4 - Neurotic Depression Anxiety depression; Depression with anxiety; Depressive reaction; Dysthymic disorder; Neurotic depressive state; Reactiv |
|  | 311 - Depressive Disorder, NEC Depressive disorder NOS; Depressive state NOS; Depression NOS |
|  | V62.8 - Other Psychological Or Physical Stress, NEC |
| Tachycardia | 785.0-Tachycardia NOS |
| Fever | 780.60-Fever NOS |
| Nausea and/or vomiting | 787.02 - Nausea alone Emesis |
|  | 787.0 - Nausea and vomiting |
|  | 787.03 - Vomiting alone Emesis |
| Chest Pain or discomfort | 786.59 - Other Chest Pain {Discomfort} {Pressure} {Tightness} in chest |
|  | 786.50 - Unspecified Chest Pain |
| Fatigue | 780.7 - Malaise And Fatigue Asthenia NOS; Lethargy; Postviral (asthenic) syndrome; Tiredness |
| Voice Disturbance | 784.49 - Other Voice Disturbance Change in voice; Dysphonia; Hoarseness; Hypernasality; Hyponasality |
| Syncope | 780.2 - Syncope And Collapse Blackout; Fainting; (Near) (Pre) syncope; Vasovagal attack |
| Sleep disturbance | 780.52 - Other Insomnia Insomnia NOS |
|  | 780.50-Sleep disturbance NOS |
| Headache | 784.0 - Headache Facial pain; Pain in head NOS |
| Runny nose and/or nasal congestion | 460 - Acute Nasopharyngitis (common Cold) Coryza (acute); Nasal catarrh, acute; Nasopharyngitis: NOS, acute, infective NOS; Rhinitis: acute, infectiv |
| Palpitation | 785.1 - Palpitations Awareness of heart beat |
| Rash | 782.1 - Rash And Other Nonspecific Skin Eruption Exanthem |
| Dyspnea and/or Shortness of breath | 786.09 - Other Dyspnea And Respiratory Abnormality Apnea; Cheyne-Stokes respiration; Respiratory: distress, insufficiency; Shortness of breath; Tachy |
| Myalgia | 729.5 - Pain In Limb |
|  | 729.1 - Myalgia And Myositis, Unspecified Fibromyositis NOS |
|  | 724.5 - Backache, Unspecified Vertebrogenic (pain) syndrome NOS |
|  | 723.1 - Cervicalgia Pain in neck |
|  | 724.2 - Lumbago Low back pain; Low back syndrome; Lumbalgia |
| Cough | 786.2 - Cough |
| Arthralgia | 719.46 - Pain In Joint Involving Lower Leg Arthralgia; Fibula; Knee joint; Patella; Tibia |
|  | 719.40 - Pain In Joint, Site Unspecified Arthralgia |
| Dizziness | 780.4 - Dizziness And Giddiness Light-headedness; Vertigo NOS |
| Diarrhea | 787.91 - Diarrhea Diarrhea NOS |
| Speech disturbance | 784.5 - Other Speech Disturbance Dysarthria; Dysphasia; Slurred speech |
| Weight loss | 783.2 - Abnormal Loss Of Weight |
| Sore throat | 784.1 - Throat Pain |
| Conjunctivitis | 372.0Acute conjunctivitis |
| General symptoms ( Amnesia / Chills / Generalized pain /Hypothermia) | 780.9 - Other General Symptoms Amnesia (retrograde); Chill(s) NOS; Generalized pain; Hypothermia, not associated with low environmental temperature |
| Abdominal pain | 789.06 - Abdominal pain, epigastric Abdominal tenderness; Colic: NOS, infantile; Cramps, abdominal; Epigastric pain; Umbilical pain |
|  | 789.00 - Abdominal pain, unspecified site Abdominal tenderness; Colic: NOS, infantile; Cramps, abdominal; Epigastric pain; Umbilical pain |
| Lymphadenopathy | 785.6 - Enlargement Of Lymph Nodes Lymphadenopathy; "Swollen glands" |
| Constipation | 564.0 - Constipation |
| Disturbance Of Skin Sensation | 782.0 - Disturbance Of Skin Sensation Anesthesia of skin; Burning or prickling sensation; Hyperesthesia; Hypoesthesia; Numbness; Paresthesia; Tinglin |
| Neuralgia | 729.2 - Neuralgia, Neuritis, And Radiculitis, Unspecified |

#### Section 3: Symptoms and time of recovery analysis

**Figure S1.**
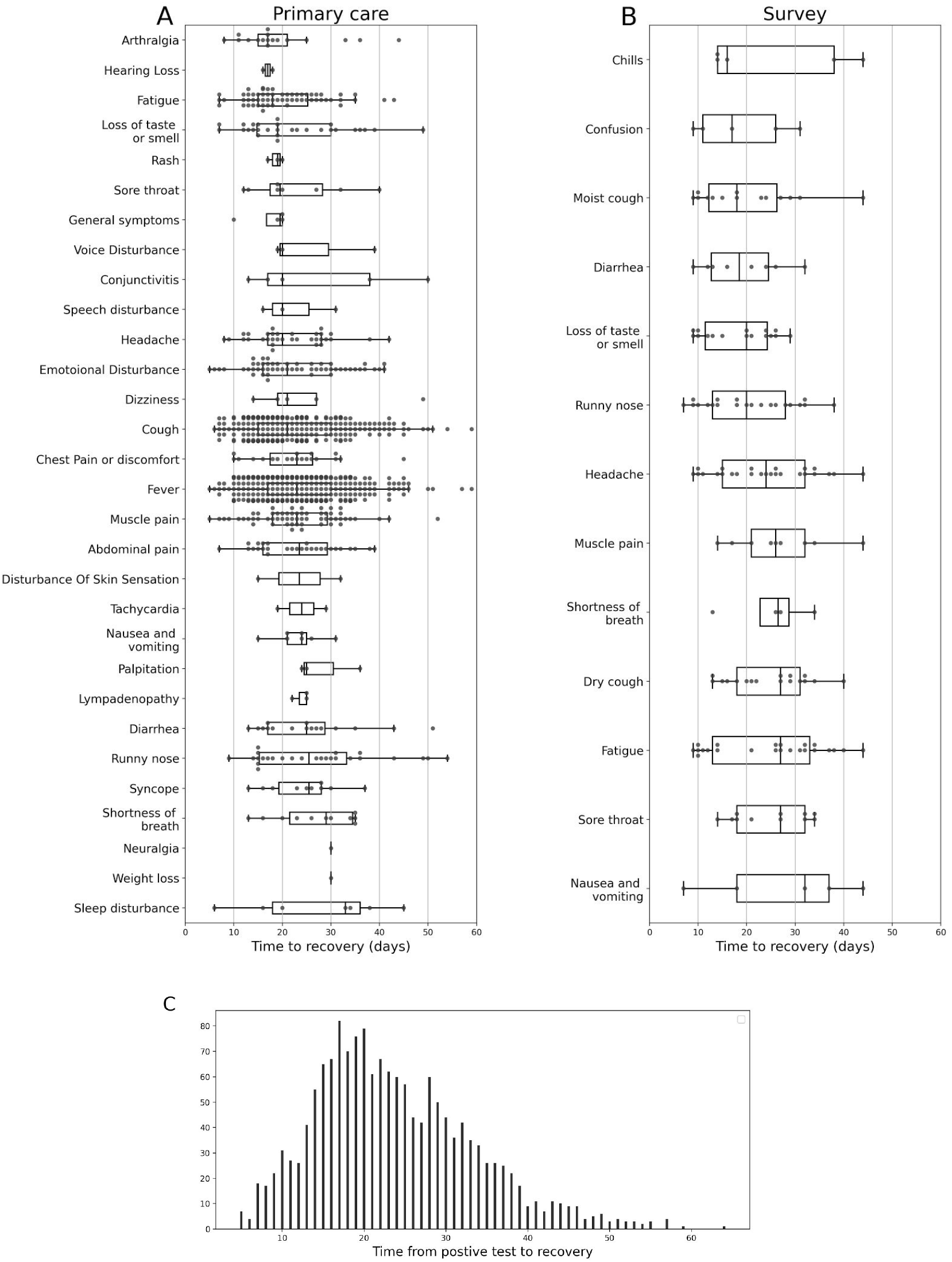
Recovery time from COVID-19. **A+B:** each point indicates recovery time for a COVID-19 positive individual who experienced a specific symptom before or after testing positive. Boxplots show quartiles of the distribution of recovery times for each symptom considered. **A** primary care visits and **B** surveys **C** Histogram presenting the number of days from diagnosis to recovery in the cohort.

#### Section 4: Odds ratio analysis

**Table S1:** Odds ratio calculation for self-reported symptoms in adults 21 days prior to the date of diagnosis. For positive COVID19 cases, this date was considered as the first positive PCR test. For COVID negative cases, this test was considered as the first negative result for COVID-19.

| Self reported symptoms in adults | Covid-19 negative who reported the symptom | Covid-19 positive who reported the symptom | COVID-19 negative w/o the symptom | COVID-19 positive w/o the symptom | OR | CI |
| --- | --- | --- | --- | --- | --- | --- |
| **Loss of taste and smell** | 110 | 18 | 4647 | 68 | **11.18** | [6.43-19.44] |
| **Confusion** | 72 | 5 | 4685 | 81 | **4.02** | [1.58-10.21] |
| **Headache** | 954 | 29 | 3803 | 57 | **2.03** | [1.29-3.19] |
| **Fatigue** | 868 | 24 | 3889 | 62 | **1.73** | [1.08-2.79] |
| Fever (Body temperature above 38°C) | 179 | 5 | 4578 | 81 | 1.58 | [0.63-3.94] |
| Dry cough | 748 | 19 | 4009 | 67 | 1.52 | [0.91-2.54] |
| Diarrhea | 317 | 8 | 4440 | 78 | 1.44 | [0.69-3.00] |
| Rhinorrhea and/or nasal congestion | 1017 | 24 | 3740 | 62 | 1.42 | [0.88-2.29] |
| Cough | 1161 | 27 | 3596 | 59 | 1.42 | [0.89-2.25] |
| Nausea and/or vomiting | 213 | 5 | 4544 | 81 | 1.32 | [0.53-3.28] |
| Wet cough | 595 | 13 | 4162 | 73 | 1.25 | [0.69-2.26] |
| Chills | 273 | 5 | 4484 | 81 | 1.01 | [0.41-2.52] |
| Myalgia | 570 | 10 | 4187 | 76 | 0.97 | [0.50-1.88] |
| Shortness of breath | 310 | 5 | 4447 | 81 | 0.89 | [0.36-2.20] |
| Other symptoms | 380 | 5 | 4377 | 81 | 0.71 | [0.29-1.77] |
| Sore throat | 908 | 12 | 3849 | 74 | 0.69 | [0.37-1.27] |
| Body temperature below 37.4 °C | 3069 | 42 | 1688 | 44 | 0.53 | [0.34-0.80] |
| Body temperature between 37.5°C and 37.9°C | 164 | 1 | 4593 | 85 | 0.33 | [0.05-2.38] |

**Table S2:** Odds ratio calculation for symptoms in children that were documented in primary care visits 21 days prior to the date of diagnosis. For positive COVID19 cases, this date was considered as the first positive PCR test. For COVID negative cases, this test was considered as the first negative result for COVID-19.

| Primary care documented symptoms in children | Covid-19 negative who reported the symptom | Covid-19 positive who reported the symptom | COVID-19 negative w/o the symptom | COVID-19 positive w/o the symptom | OR | CI |
| --- | --- | --- | --- | --- | --- | --- |
| Disturbances Of Sensation Of Smell And Taste | 12 | 1 | 17140 | 583 | 2.45 | [0.32-18.87] |
| Syncope | 24 | 2 | 17128 | 582 | 2.45 | [0.58-10.40] |
| Emotional Disturbance | 131 | 9 | 17021 | 575 | 2.03 | [1.03-4.02] |
| Sleep disturbance | 20 | 1 | 17132 | 583 | 1.47 | [0.20-10.97] |
| Fatigue | 212 | 10 | 16940 | 574 | 1.39 | [0.73-2.64] |
| Dizziness | 25 | 1 | 17127 | 583 | 1.18 | [0.16-8.69] |
| Conjunctivitis | 102 | 4 | 17050 | 580 | 1.15 | [0.42-3.14] |
| Rash | 144 | 4 | 17008 | 580 | 0.81 | [0.30-2.21] |
| Sore throat | 177 | 4 | 16975 | 580 | 0.66 | [0.24-1.79] |
| Myalgia | 137 | 3 | 17015 | 581 | 0.64 | [0.20-2.02] |
| Arthralgia | 48 | 1 | 17104 | 583 | 0.61 | [0.08-4.44] |
| Headache | 211 | 4 | 16941 | 580 | 0.55 | [0.21-1.49] |
| Runny nose and or nasal congestion | 125 | 2 | 17027 | 582 | 0.47 | [0.12-1.90] |
| Speech disturbance | 65 | 1 | 17087 | 583 | 0.45 | [0.06-3.26] |
| Dyspnea and or Shortness of breath | 269 | 4 | 16883 | 580 | 0.43 | [0.16-1.17] |
| Cough | 1965 | 29 | 15187 | 555 | 0.4 | [0.28-0.59] |
| Fever | 3305 | 39 | 13847 | 545 | 0.3 | [0.22-0.42] |
| Nausea and or vomiting | 131 | 1 | 17021 | 583 | 0.22 | [0.03-1.60] |
| Diarrhea | 332 | 2 | 16820 | 582 | 0.17 | [0.04-0.70] |
| Abdominal pain | 299 | 1 | 16853 | 583 | 0.1 | [0.01-0.69] |

**Table S3:** Odds ratio calculation for symptoms in Adults that were documented in primary care visits 21 days prior to the date of diagnosis. For positive COVID19 cases, this date was considered as the first positive PCR test. For COVID negative cases, this test was considered as the first negative result for COVID-19.

| Primary care documented Symptoms in Adults | Covid-19 negative who reported the symptom | Covid-19 positive who reported the symptom | COVID-19 negative w/o the symptom | COVID-19 positive w/o the symptom | OR | CI |
| --- | --- | --- | --- | --- | --- | --- |
| **Disturbances Of Sensation Of Smell And Taste** | 137 | 31 | 67638 | 2800 | **5.47** | [3.69-8.09] |
| **Runny nose and or nasal congestion** | 404 | 35 | 67371 | 2796 | **2.09** | [1.47-2.95] |
| **Syncope** | 126 | 11 | 67649 | 2820 | **2.09** | [1.13-3.88] |
| **Fever** | 5157 | 334 | 62618 | 2497 | **1.62** | [1.44-1.83] |
| Speech disturbance | 35 | 2 | 67740 | 2829 | 1.37 | [0.33-5.69] |
| Lymphadenopathy | 86 | 4 | 67689 | 2827 | 1.11 | [0.41-3.04] |
| Sleep disturbance | 228 | 9 | 67547 | 2822 | 0.94 | [0.48-1.84] |
| Voice Disturbance | 76 | 3 | 67699 | 2828 | 0.94 | [0.30-3.00] |
| Cough | 7202 | 271 | 60573 | 2560 | 0.89 | [0.78-1.01] |
| Fatigue | 1748 | 65 | 66027 | 2766 | 0.89 | [0.69-1.14] |
| Tachycardia | 59 | 2 | 67716 | 2829 | 0.81 | [0.20-3.32] |
| Headache | 993 | 33 | 66782 | 2798 | 0.79 | [0.56-1.12] |
| General symptoms ( Amnesia<br>Chills Generalized pain<br>Hypothermia) | 156 | 5 | 67619 | 2826 | 0.77 | [0.31-1.87] |
| Nausea and or vomiting | 346 | 11 | 67429 | 2820 | 0.76 | [0.42-1.39] |
| Myalgia | 2613 | 76 | 65162 | 2755 | 0.69 | [0.55-0.87] |
| Emotional Disturbance | 2331 | 66 | 65444 | 2765 | 0.67 | [0.52-0.86] |
| Arthralgia | 945 | 26 | 66830 | 2805 | 0.66 | [0.44-0.97] |
| Dizziness | 420 | 10 | 67355 | 2821 | 0.57 | [0.30-1.07] |
| Palpitation | 215 | 5 | 67560 | 2826 | 0.56 | [0.23-1.35] |
| Abdominal pain | 1519 | 34 | 66256 | 2797 | 0.53 | [0.38-0.75] |
| Dyspnea and or Shortness of breath | 425 | 9 | 67350 | 2822 | 0.51 | [0.26-0.98] |
| Chest Pain or discomfort | 1185 | 25 | 66590 | 2806 | 0.5 | [0.34-0.75] |
| Hearing Loss | 153 | 3 | 67622 | 2828 | 0.47 | [0.15-1.47] |
| Diarrhea | 921 | 18 | 66854 | 2813 | 0.46 | [0.29-0.74] |
| Sore throat | 672 | 10 | 67103 | 2821 | 0.35 | [0.19-0.66] |
| Weight loss | 146 | 2 | 67629 | 2829 | 0.33 | [0.08-1.32] |
| Disturbance Of Skin Sensation | 149 | 2 | 67626 | 2829 | 0.32 | [0.08-1.30] |
| Neuralgia | 84 | 1 | 67691 | 2830 | 0.28 | [0.04-2.05] |
| Conjunctivitis | 179 | 1 | 67596 | 2830 | 0.13 | [0.02-0.95] |

## Notes

### Competing Interest Statement

The authors have declared no competing interest.

### Funding Statement

No funding was received for this article

### Author Declarations

The study protocol was approved by Maccabi Health Services' institutional review board (0024-20-MHS)

